# *In vivo* PET imaging of neuroinflammation in familial frontotemporal dementia

**DOI:** 10.1101/2020.05.11.20098046

**Authors:** Maura Malpetti, Timothy Rittman, P. Simon Jones, Thomas E. Cope, Luca Passamonti, W. Richard Bevan-Jones, Karalyn Patterson, Tim D. Fryer, Young T. Hong, Franklin I. Aigbirhio, John T. O’Brien, James B. Rowe

**Author notes:** Joint Senior authors (John T. O’Brien, James B. Rowe). **Corresponding Author:** Prof. James B. Rowe, Department of Clinical Neurosciences, University of Cambridge, Herchel Smith Building, Forvie Site, Robinson Way, Cambridge Biomedical Campus, Cambridge CB2 0SZ.

## Abstract

**INTRODUCTION:** We report *in vivo* patterns of neuroinflammation and abnormal protein aggregation in seven cases of familial frontotemporal dementia with mutations in MAPT, GRN and C9orf72 genes.

**METHODS:** Using positron emission tomography (PET), we explored the association of the distribution of activated microglia, as measured by the radioligand [^11^C]PK11195, and the regional distribution of tau- or TDP-43 pathology, indexed using the radioligand [^18^F]AV-1451. The familial FTD PET data were compared to healthy controls.

**RESULTS:** Familial FTD patients across all mutation groups showed increased [^11^C]PK11195 binding predominantly in frontotemporal regions, with additional regions showing abnormalities in individuals. Patients with MAPT mutations had a consistent distribution of [^18^F]AV-1451 binding across the brain, with heterogeneous distributions among carriers of GRN and C9orf72 mutations.

**DISCUSSION:** This case series suggests a consistent role for neuroinflammation in the pathophysiology of familial FTD, warranting further consideration of immunomodulatory therapies for disease modification and prevention.

## 1. INTRODUCTION

A fifth of frontotemporal dementia (FTD) cases are autosomal dominant[1], most commonly arising from mutations in MAPT, GRN or C9orf72 genes[2]. The pathological and clinical features of familial FTD closely resemble sporadic cases, but mutation carriers allow inference of the underlying pathology with molecular specificity. Although misfolding and aggregation of tau or TDP-43 protein leads to characteristic FTD neuropathology, neuroinflammation may be an early etiopathogenetic process, rather than a consequence of neurodegeneration[3-8]. Positron emission tomography (PET) with the radioligand [^11^C]PK11195 reveals increased neuroinflammation in frontotemporal regions in FTD[5,6], with neuroinflammation preceding the onset of symptoms[3,4]. However, the pattern of inflammation by genotype, and the relationship to phenotype and protein aggregates, is unknown.

We used [^11^C]PK11195 to quantify *in vivo* neuroinflammation in patients with symptomatic familial FTD. We then compared the distribution of inflammation by phenotype, and to the distribution of tau or TDP-43 using [^18^F]AV-1451, a radioligand sensitive to tau and TDP-43 pathology.

## 2. METHODS

Seven patients with familial FTD were recruited from the Cambridge University Centre for Frontotemporal Dementia: 2 with MAPT 10+16 gene mutation, 2 with GRN C388_391delCAGTp.(Gln130fs) and 3 with C9orf72 expansions. Six patients met diagnostic criteria for behavioural variant FTD (bvFTD)[9] and one for non-fluent variant primary progressive aphasia (nfvPPA)[10]. Six were included alongside 25 sporadic FTD cases in a previous publication using different analyses[6]. They underwent a structured clinical and neuropsychological assessment, together with brain imaging with 3T MRI, [^11^C]PK11195 PET and [^18^F]AV-1451 PET [11]. The study was approved by the National Research Ethic Service’s East of England Cambridge Central Committee. Written informed consent was obtained from all participants.

Structural MRI, [^11^C]PK11195 PET and [^18^F]AV-1451 PET data were acquired and processed as previously described [6,11]. To quantify the density of radioligand binding, non-displaceable binding potential (BP_ND_) was calculated for both [^11^C]PK11195 and [^18^F]AV-1451 at the voxel level and also in 83 regions of interest (ROIs) based on a modified version of the n30r87 Hammersmith atlas (www.brain-development.org), with ROI data corrected for cerebrospinal fluid (CSF) partial volume prior to kinetic modeling. In view of substantial off-target subcortical binding, we confine our analyses to cortical regions.

One-tailed Crawford tests for single-case analysis[12] were performed on regional values for each radioligand, comparing each patient with an age-, sex-matched group of healthy elderly adults (Supplementary Table 1), who underwent either [^11^C]PK11195 PET (N=15, Supplementary Table 2) or [^18^F]AV-1451 PET (N=15, Supplementary Table 3).

## 3. RESULTS

Individual BP_nd_ maps for [^11^C]PK11195 and [^18^F]AV-1451 are reported in Figures 1.

**Figure 1.**
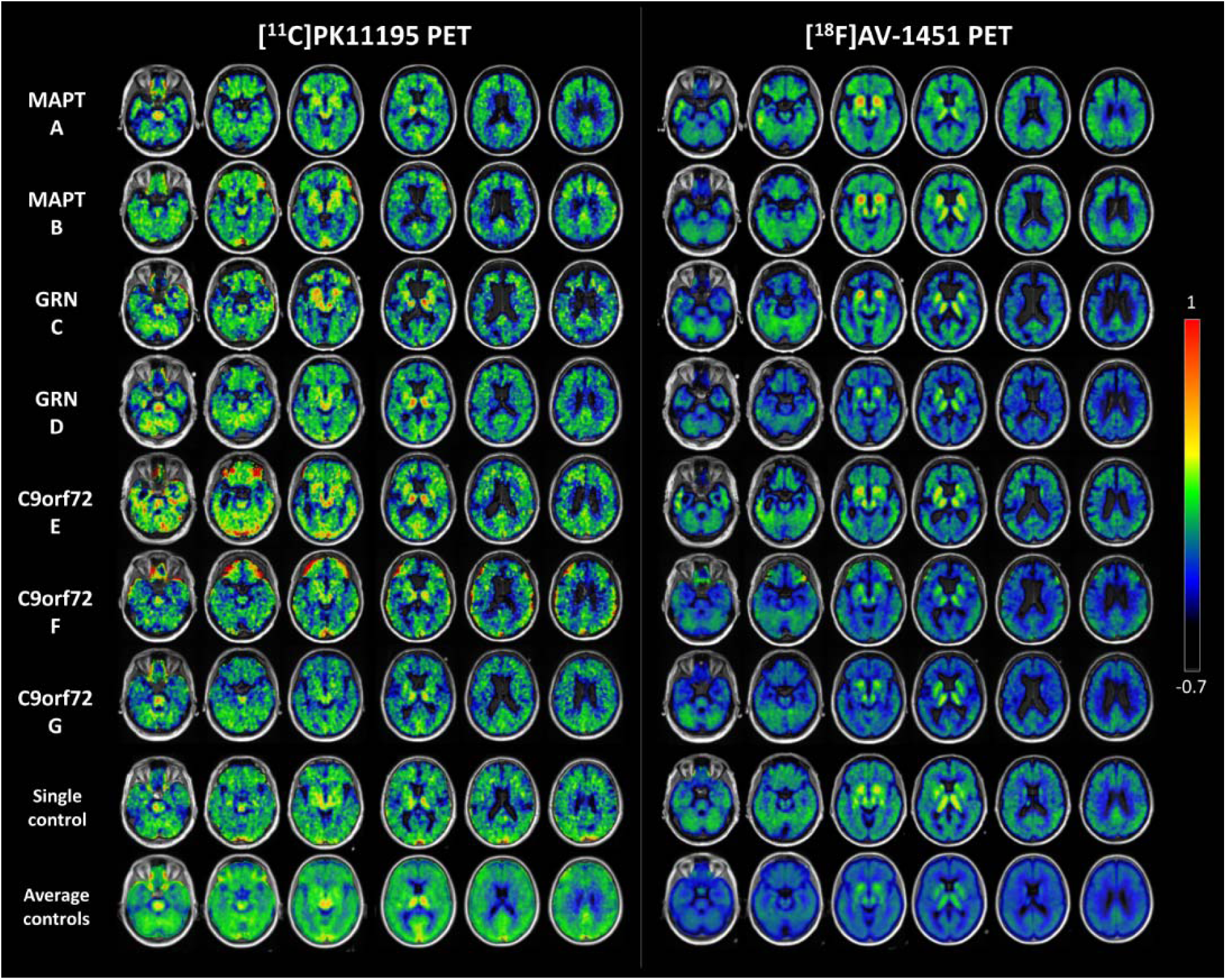
Axial slices of the [^11^C]PK1195 (left panel) and [^18^F]AV-1451 BP_ND_ (right panel) binding potential maps for each patient (A-G). The last two rows of each panel represent the ligand-specific maps of a single control and average BP_ND_ images across 15 controls. Patients A and B are MAPT mutation carriers; cases C and D are patients with GRN mutations; and patients E, F and G are C9orf72 mutation carriers. BP_ND_ scale bar runs along the right side of the figure. The slices are aligned in MNI space.

### MAPT mutations

*Patient A* was diagnosed with bvFTD aged 51, with progressive lack of insight and language, impairment of memory and judgement, loss of functional skills, apathy, and loss of semantic knowledge. She scored 43/100 on the Addenbrooke’s Cognitive Examination Revised (ACE-R), 9/18 on the Frontal Assessment Battery (FAB), 2/30 on the on Frontotemporal Dementia Rating Scale (FRS), and 118/180 on the Cambridge Behavioural Inventory Revised (CBI-R). She had a family history of FTD, with her father carrying a symptomatic MAPT 10+16 mutation. [^11^C]PK11195 binding was elevated in the lateral temporal lobes and temporal poles, dorsolateral and orbitofrontal cortex, posterior cingulum, and inferior parietal cortex. Elevated [^18^F]AV-1451 binding overlapped with [^11^C]PK11195, in the anterior and lateral temporal lobes, dorsolateral and orbitofrontal cortex, posterior cingulum and inferior parietal lobules.

*Patient B* presented at 60 years of age with bvFTD, characterised by obsessional and compulsive behaviours, comprehension impairments, anomia, and a positive family history of dementia consistent with FTD. She scored 44/100 on ACE-R, 7/18 on FAB, 1/30 on FRS and 99/180 on CBI-R. PET scans showed increased [^11^C]PK11195 binding in the frontal lobes (particularly the dorsolateral prefrontal cortex), medial temporal lobe, angular gyrus, and posterior cingulum. The [^18^F]AV-1451 binding pattern was similar to [^11^C]PK11195, in the frontal lobes, most prominently in the dorsolateral prefrontal and orbitofrontal cortex, angular gyrus, and posterior cingulum, with only mildly increased temporal binding.

### GRN mutations

*Patient C* presented bvFTD at age 70 with apathy, emotional incontinence and disinhibition, but no motor signs. She scored 33/100 on ACE-R, 3/30 on FRS, and 88/180 on CBI-R. [^11^C]PK11195 binding was patchily increased in the temporal poles, insular cortex and orbitofrontal cortex. [^18^F]AV-1451 PET showed mildly increased binding in the lateral temporal regions, posterior cingulum, and cerebellum.

*Patient D* presented progressive non-fluent aphasia at age 65 with additional mild behavioural changes including jocularity and hoarding of sweets. He was diagnosed with nfvPPA 18 months after his symptom onset. By the time of the PET scans, he had declined significantly with minimal speech, yes/no confusion and significant behavioural problems without motor involvement. He scored 76/100 on ACE-R, 9/18 on FAB, 12/30 on FRS, and 62/180 on CBI-R. There was no significant binding of [^18^F]AV-1451, while [^11^C]PK11195 binding was increased in the cerebellum and anterior temporal lobes.

### C9orf72 mutations

*Patient E* presented bvFTD aged 56 years, with irritable behaviour, over-valued ideas and obsessions, a sweet tooth, semantic impairment, but no signs of motor neuron disease. He scored 53/100 on ACE-R, 6/18 on FAB, 9/30 on FRS, and 76/180 on CBI-R. On PET imaging there was increased [^11^C]PK11195 binding in the anterior temporal lobes, orbitofrontal and temporal regions, and cerebellum. There was increased [^18^F]AV-1451 binding in temporal lobes, most notably in the lateral regions, and in the superior frontal cortex.

*Patient F* presented bvFTD at age 51 with behavioural changes, abnormal beliefs, stereotypical and repetitive behaviours, but no features of motor neuron disease. She developed paranoid delusions, apathy, a sweet tooth with significant weight gain, and a moderate aphasia with non-fluent speech and anomia. She scored 45/100 on ACE-R, 7/18 on FAB, 3/30 on FRS, and 60/180 on CBI-R. Her pattern of [^18^F]AV-1451 binding was low across the whole-brain, and only very mild excess binding in the lateral orbitofrontal and inferior parietal regions. However, [^11^C]PK11195 PET showed increased binding across frontotemporal and parietal regions, with peaks in the orbitofrontal and anterior temporal regions.

*Patient G* presented bvFTD at age 58 with a long history of psychiatric symptoms and an episode of psychotic depression at age 44 following bereavements. At age 56, he had a relapsing episode of depression with psychotic features and he was consequently treated with risperidone after which he developed a parkinsonian syndrome. His psychosis progressed and he developed disinhibited social behaviours and memory problems. There was a positive family history for motor neuron disease, although he had no signs or symptoms of motor neuron diseases. He scored 46/100 on ACE-R, 5/18 on FAB, 2/30 on FRS, and next of kin endorsed 140/180 items on CBI-R. [^11^C]PK11195 PET showed increased binding in parietal regions, posterior cingulum, precuneus and cerebellum. There was mildly increased [^18^F]AV-1451 binding in parietal-occipital regions, posterior cingulate cortex, and cerebellum.

## 4. DISCUSSION

The study demonstrates *in vivo* neuroinflammation with the three most common monogenic forms of FTD. Familial FTD was consistently associated with neuroinflammation in frontotemporal regions, and often parietal cortex, across all genes. The distribution of neuroinflammation reflected clinical heterogeneity, with underlying tau or TDP-43 pathology. In both MAPT patients, the pattern of [^11^C]PK11195 binding overlapped with [^18^F]AV-1451 binding distribution across the brain, with particular elevation in frontotemporal and parietal regions. The GRN cases had different phenotypes which may reflect the variable pathology distribution. The distribution of suprathreshold neuroinflammation was wider than [^18^F]AV-1451 binding, being especially increased in anterior temporal regions in both patients, and orbitofrontal cortex in patient C. C9orf72 carriers presented a notable neuroinflammation pattern involving frontotemporal and parietal regions, which aligned with anterior-posterior MRI changes associated with this mutation [13]. In these patients, who are expected to have TDP-43 pathology but not significant tauopathy, [^18^F]AV-1451 binding was not intense or extensive, compared to the distribution of neuroinflammation. [^18^F]AV-1451 was less marked than in the alternative TPD-43 pathology subtype associated with semantic dementia[14].

These results emphasise the role of neuroinflammation in mediating the pathophysiology of genetic FTD and its associated clinical syndromes[15]. In patients with MAPT mutations, neuroinflammation may interact with the misfolding and aggregation of mutant tau, while preclinical models of C9orf72 and GRN cases, neuroinflammation can be triggered by dipeptide repeats or progranulin haploinsufficiency in advance of the large TDP-43 aggregates.

We suggest that future disease-modifying treatment strategies may be enhanced by immunomodulation. [^11^C]PK11195 PET studies in pre-symptomatic carriers could help develop early, targeted, interventions.

## Data Availability

Anonymized data may be shared by request to the senior author from a qualified investigator for non-commercial use (sharing of some data is subject to restrictions according to participant consent and data protection legislation).

## Acknowledgments

We thank our patients and their families for their participation in this study. We also thank radiographers/technologists at the Wolfson Brain Imaging Centre and Addenbrooke’s PET/CT Unit, and the research nurses of the Cambridge Centre for Frontotemporal Dementia and Related Disorders for their invaluable support in data acquisition. We thank the East Anglia Dementias and Neurodegenerative Diseases Research Network (DeNDRoN) for help with subject recruitment, and Drs Istvan Boros, Joong-Hyun Chun, and other WBIC RPU staff for the manufacture of the radioligands. We thank Avid (Lilly) for supplying the precursor for the production of [^18^F]AV-1451 used in this study.

## Competing interests

JBR reports consultancy unrelated to the work with Biogen, UCB, Asceneuron and Althira; and receipt of research grants from Janssen, AZ-Medimmune, Lilly unrelated to this work. JOB has provided consultancy to TauRx, Axon, Roche, GE Healthcare, and Lilly; and has research awards from Alliance Medical and Merck. TR has received honoraria from the National Institute for Health and Clinical Excellence, Oxford Biomedica and Learna Ltd.

## Funding

This study was funded by the NIHR Cambridge Biomedical Research Centre: Dementia and Neurodegeneration theme; the Wellcome Trust (103838); a Cambridge Trust & Sidney Sussex College Scholarship; the Medical Research Council; and the Cambridge Centre for Parkinson-Plus.

## Notes

### Competing Interest Statement

The authors have declared no competing interest.

